# Assessing the impact of preventive mass vaccination campaigns on yellow fever outbreaks in Africa : a population-level self-controlled case-series study

**DOI:** 10.1101/2020.07.09.20147355

**Authors:** Kévin Jean, Hanaya Raad, Katy A. M. Gaythorpe, Arran Hamlet, Judith E. Mueller, Dan Hogan, Tewodaj Mengistu, Heather J. Whitaker, Tini Garske, Mounia N. Hocine

## Abstract

**Introduction:** The Eliminate Yellow fever Epidemics (EYE) strategy was launched in 2017 in response to the resurgence of yellow fever in Africa and the Americas. The strategy relies on several vaccination activities, including preventive mass vaccination campaigns (PMVCs). However, by how much PMVCs are associated with a decreased risk of outbreak to occur has not yet been quantified.

**Methods and Findings:** We used the self-controlled case series (SCCS) method to assess the association between the occurrence of yellow fever outbreaks and the implementation of PMVCs at the province level in the African endemic region. As all time-invariant confounders are implicitly controlled for, the SCCS method is an alternative to classical cohort or case-control study designs when the risk of residual confounding is high, in particular confounding by indication. The location and dates of outbreaks were identified from international epidemiological records, and information on PMVCs was provided by coordinators of vaccination activities and international funders. The study sample consisted of provinces that were both affected by an outbreak and targeted for a PMVC between 2005 and 2018. We compared the relative incidence of outbreaks before and after the implementation of a PMVC. The sensitivity of our estimates to a range of assumptions was explored, and the results of the SCCS method were compared to those obtained through a retrospective cohort study design. We further derived the number of yellow fever outbreaks that have been prevented by PMVCs.

The study sample consisted of 33 provinces from 11 African countries. Among these, outbreaks occurred during the pre-PMVC period in 26 (79%) provinces versus 7 (21%) occurring in the post-PMVC period. At the province level, the post-PMVC period was associated with a 86% reduction (95% Confidence interval 66% to 94%, p-value<0.001) in the risk of outbreak as compared to the pre-PMVC period. This negative association was robustly observed across a range of sensitivity analyses, especially when using quantitative estimates of vaccination coverage as alternative exposure, or when varying the observation period. Conversely, the results of the cohort-style analyses were highly sensitive to the choice of covariates included in the model. Based on the SCCS results, we estimated that PMVCs were associated with a 34% (22% to 45%) reduction in the number of outbreaks in Africa over the study period.

A limitation of our study is the fact that it does not account for potential time-varying confounders, such as changing environmental drivers of yellow fever or possibly improved disease surveillance.

**Conclusion:** In this study, we provide new empirical evidence of the high preventive impact of PMVCs on yellow fever outbreaks. This study illustrates that the SCCS method can be advantageously applied at the population level in order to evaluate a public health intervention.

**AUTHOR SUMMARY:** *Why Was This Study Done?:*

- Yellow fever is a mosquito-borne vaccine-preventable disease that may cause large urban outbreaks, especially in tropical African regions.
- Since 2006, Preventive Mass Vaccination Campaigns (PMVCs) have been implemented in many African provinces. These are large scale vaccination campaigns targeting all or most age group in a specific area.
- The preventive impact PMVCs may have on the risk of yellow fever outbreak has not been quantified yet.

*What Did the Researchers Do and Find?:*

- We used the self-controlled case series (SCCS) method to assess the association between PMVCs and outbreak risk at the province level over 34 African countries between 2005 and 2018.
- Using this method, we compared pre- and post-PMVC period are compared within each province individually, thus controlling for all possible confounders that do not vary in time, especially the fact that provinces indicated for PMVCs are generally those considered at highest baseline risk of yellow fever (ie confounding by indication).
- At the province level, we estimated that implementation of PMVCs was associated with an 86% reduction (66% to 94%) in the risk of yellow fever outbreak.
- Results from a complete cohort analysis provided less reliable results than the SCCS method, likely because of confounding by indication that was not entirely controlled for by adjusting on known drivers of yellow fever.
- We further estimated that all PMVCs conducted between 2006 and 2018 in Africa may have reduced the total number of yellow fever outbreaks by 34% (22% to 45%).

*What Do These Findings Mean?:*

- These results provide new empirical evidence of the high preventive impact of PMVCs on yellow fever outbreaks.
- These results may encourage rapid re-scheduling of yellow fever PMVCs that have been postponed due to the COVID-19 pandemic.
- The SCCS design may be advantageously applied at the population level to assess the impact of public health interventions.

## INTRODUCTION

Recent years have seen the resurgence of Yellow fever outbreaks in Africa and Latin America [1]. Regarding Africa specifically, five alerts have been issued for the first semester 2020 alone (Uganda, South Sudan, Ethiopia, Togo, Gabon) [2]. As a response to the large-scale Angola 2015-2016 outbreak, the World Health Organization (WHO) launched the Eliminate Yellow fever Epidemics (EYE) initiative in 2017 [3]. This strategy aims at preventing sporadic cases sparking urban outbreaks and potentially triggering international spread. It relies on various vaccination activities, including Preventive Mass Vaccination Campaigns (PMVCs) that target all or most age groups in a specific area. Evaluating the health impact of such campaigns is key to inform further PMVCs within or beyond the EYE strategy, to ensure population acceptance and adherence to vaccination campaigns, and to sustain domestic and international efforts for vaccination activities.

Previous attempts were made in order to estimate the impact of vaccination activities, including PMVCs [4–6]. These attempts mostly relied on mathematical models to estimate PMVCs impact in terms of deaths or cases prevented on the long term. However, few studies aimed at quantifying the effect of vaccination campaigns on the risk of outbreak. Regardless of the number of cases they may generate, outbreaks can possibly lead to healthcare, economic and social destabilizations of entire regions. As an example, the west-African 2013-2016 Ebola outbreak strained health systems and generated fear of the disease. This caused excess deaths due to neglected need for malaria control [7,8].

When assessed at the population level, the association of vaccination activities and risk of outbreak can be approached within a classical epidemiological perspective. As individuals would be in a cohort study, populations (for instance populations living in well-defined geographical areas) may be followed over time while tracking both exposure (vaccination activities) and events (outbreaks). In such observational studies, a risk of confounding arises when both exposure and event share a same cause. This risk is high when measuring the association between PMVCs and yellow fever outbreaks because PMVCs usually target areas that are assessed at particularly high risk by public health officials, due to the disease circulation in the past or based on expert view or risk assessment [9]. Such a risk of confounding is usually overcome in the statistical analysis by conditioning, generally adjusting, on the shared common cause; in this case the baseline risk of yellow fever in the area. However, the environmental or demographic drivers of yellow fever are not fully understood [10,11], leading to a situation in which residual confounders may bias the measure of association.

The self-controlled case series (SCCS) method is a case-only epidemiological study design for which individuals are used as their own control [12]. As all known and unknown time-invariant confounding are implicitly controlled for, the method is a relevant alternative to classical cohort or case-control study designs when the risk of residual confounding is high. The SCCS method has successfully been applied at the individual level, comparing exposure vs. non-exposure periods within individual cases [13]. However, to our knowledge, this method has never been used for population-level case series to evaluate the health effects of a public health intervention in specific regions, countries, or other predefined geographical clusters that may be considered as group-level cases.

Here, we illustrate the use of the SCCS method at the population level by assessing the association between the implementation of PMVCs and the occurrence of yellow fever outbreaks at the province level in the African endemic region between 2005 and 2018.

## METHODS

### Study hypotheses

Considering the yellow fever vaccine’s high level of efficacy [14], and given the fact that PMVCs target all or most age groups in targeted areas, we expect to detect a substantial preventive effect of PMVC on the risk of outbreak. We also expect to detect this association in a cohort design, providing confounders in the association between exposure to PMVC and outbreak are adequately controlled for (no model misspecification). A SCCS model would avoid the risk of residual confounding, at least for time-independent variables, but would reduce statistical power as compared to a cohort-design analysis [15].

### Data used

This study relied on datasets that were previously collected and regularly updated for a broader project aiming at estimating the burden of yellow fever and the impact of vaccine activities [4,6]. The analytic plan was defined before the start of the analysis. This study did not require ethical approval.

In 2005, the African Regional Office of WHO established a yellow fever surveillance database across 21 countries in West and Central Africa based on reports of suspected yellow fever cases [4]. This is likely to have influenced the standards of yellow fever surveillance in these countries. In order to reduce the possible effect of time-changing surveillance quality, the beginning of the study period was set at this date. We compiled location and dates of yellow fever outbreaks reported in Africa between 2005 and 2018 from international epidemiological reports, namely the WHO Weekly Epidemiological Reports (WER) and the Diseases Outbreak News (DON) [16,17]. As per WHO recommendations, a single, confirmed yellow fever case is sufficient to justify outbreak investigation. Based on the results of the investigation and on the absence of other suspected cases, an alert can be classified as an isolated case, which was not considered for this study [18]. Locations were resolved at the first sub-national administrative level, thereafter called province, and data were recorded for each outbreak with the date of occurrence. Outbreak reports that could not be located at the province level were excluded.

We compiled data regarding PMVCs conducted as part of the Yellow Fever initiative since 2006 [19], and additional campaigns further conducted under the EYE strategy [1]. Starting dates and locations of PMVCs were collected, and the resulting list of vaccination campaigns was compared with data from the WHO International Coordinating Group (ICG) on Vaccine Provision, while resolving any discrepancy. This virtually ensured completeness of information regarding mass vaccination activities. Note that information on the vaccine strain used for each PMVCs are not widely available, however, both vaccine strains recommended by the WHO (17D and 17DD) do not differ in terms of immunogenicity [14,20]. PMVCs considered here implied full-dose vaccine only as fractional-dose vaccination has been approved by WHO only as part of an emergency response to an outbreak if there is a shortage of full-dose YF vaccine [21].

Estimates of population-level vaccine-induced protection against yellow fever were obtained from Hamlet et al [22]. These estimates were obtained by compiling regularly updated vaccination data from different sources (routine infant vaccination, reactive campaigns, PMVCs) and inputting these into a demographic model.

#### Main SCCS analysis

For our main analysis, we used the SCCS method to estimate the incidence rate ratio (IRR) of yellow fever outbreak after vs. before the implementation of a PMVC. We used the province as unit of analysis, so that the main outcome represents the risk for a province to be affected by an outbreak. As the dependency between potential outbreak recurrences in the same province could not be excluded, we limited the analyses to the first outbreak occurrence per province for the main analysis [23]. We used a conditional Poisson model with logit link to model the occurrence of outbreaks [15].

Provinces included in the SCCS analysis were those both affected by an outbreak and targeted for a PMVC over a study period from Jan 1^st^ 2005 to Dec 31^st^ 2018. We defined the unexposed period as the pre-PMVC period. Previous research found that a single dose of yellow fever vaccine provides a long-lasting immunity with high efficacy [14,20]. Therefore, and given the relatively short observation period of the study (14 years), we assumed the exposure period started at the date of the first PMVC and lasted until the end of the observation period. This assumption was made regardless of estimated achieved coverage or intra-province geographic extent of the campaigns. In other terms, we assumed the campaigns achieved uniform high coverage in all age groups across provinces and that this high coverage was maintained up to the end of the study period. This assumption of persisting high coverage is further justified by the inclusion of yellow fever vaccine in the Enhanced Program of Immunization of most of the countries in the study region [24]. Routine infant vaccination may not rapidly increase population-level immunity by itself, but does contribute to maintain high levels once achieved by the means of PMVCs.

#### Alternative SCCS models and sensitivity analyses

In order to allow for possible variation in coverages achieved across PMVCs, we considered the estimated population-level vaccine coverage as an alternative time-varying, quantitative exposure (considered as categories with 20% bandwidth: 0-19% / 20-39% / 40-59% / 60-79% / 80-100%).

We also used alternative SCCS models to assess the influence of several assumptions on our results (Table 1) [12]. We conducted a SCCS analysis considering all outbreaks, instead of the first one only, in order to evaluate the influence of the assumption of non-independent recurrence. Additionally, as it is possible that the occurrence of an outbreak could affect subsequent exposure, we conducted a SCCS analysis including a 3-year pre-exposure period.

**Table 1:** Analysis plan for the measure of the association between yellow fever vaccination activities and outbreak risk. SCCS: Self-controlled case series.

| Model | Outcome |  | Exposure |  |  | Covariates |  |  |
| --- | --- | --- | --- | --- | --- | --- | --- | --- |
|  | 1<sup>st</sup> outbreak only | Repeated outbreaks included | Binary exposure: pre- vs. post-PMVC | Inclusion of a 3-year pre-exposure window period | Quantitative estimates of population-level vaccination coverage | None (self-controlled) | Covariates used in a previous statistical model | Covariates used in a previous mechanistic model |
| SCCS Model 1 (main analysis) | X |  | X |  |  | X |  |  |
| SCCS Model 2 |  | X | X |  |  | X |  |  |
| SCCS Model 3 | X |  |  | X |  | X |  |  |
| SCCS Model 4 | X |  |  |  | X | X |  |  |
| Cohort model 1 | X |  | X |  |  |  | X |  |
| Cohort model 2 | X |  |  |  | X |  | X |  |
| Cohort model 3 | X |  | X |  |  |  |  | X |
| Cohort model 4 | X |  |  |  | X |  |  | X |

PMVC: preventive mass vaccination campaign.

As the precise date of outbreaks and PMVCs start were not always available, we assumed where missing, that outbreaks started in the middle of the year and that exposure to PMVCs started at the end of the year. The influence of these assumptions was explored in sensitivity analysis.

Furthermore, to assess how the choice of the study period influenced our results, we also conducted a sensitivity analysis considering alternative start and end dates: i) Jan 1^st^ 2007 to Dec 31^st^ 2018, and ii) Jan 1^st^ 2005 to Dec 31^st^ 2014.

Lastly, to assess whether spatial autocorrelation could affect our results, we conducted multiple re-sampling. In each re-sampling from the SCCS study sample, we only sampled one random province per country and re-estimated the IRR of the association between exposure and the event. This implicitly accounts for spatial autocorrelation within, but not across countries.

#### Analysis using the cohort design

We compared the results obtained using the SCCS method with those obtained using a classical cohort design. The study sample was constituted of all provinces belonging to the 34 African countries at high or moderate risk for yellow fever [3]. We used univariate and multivariate Poisson regression models with robust variance, considering exposure alternatively as a binary (pre-versus post-PMVC) or continuous (vaccination coverage) time-dependent variable.

In a cohort design, the choice of covariates to include is critical to prevent bias due to residual confounding. However, there is currently no clear consensus about the demographical and environmental drivers of yellow fever. We thus considered two (partially overlapping) sets of covariates (Supplementary Text S1). Both sets of variables were documented to reproduce well the presence and absence of yellow fever records at the province level. The first set of covariates was previously used in a statistical model whereas the second was used in a mechanistic model.[4,10] Statistical models aim to describe the patterns of association between species (including infectious agents species) and environmental variables while mechanistic models aim at explicitly representing biological processes in their occurrence [25]. The association between each covariate and the exposure status was explored using modified Poisson regression.

### Number of outbreaks averted and prevented fraction

For each province,, we estimated the expected number of outbreaks averted by PMVC, *A*_*i*_, using the formula:

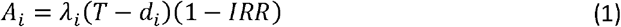

Where *T* is the total time of observation, *d*_*i*_ is the time at which PMVC was implemented (if no PMVC in province *i*, thus *d*_*i*_ *= T*),, *λ* _*i*_ is the rate of outbreak occurrence in a Poisson process in the absence of PMVC, and IRR is the incidence rate ratio after vs. before PMVC implementation. With 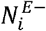 being the number of outbreaks observed in the province *i* during the pre-PMVC, an estimator of *λ* _*i*_ is 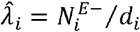, which leads to:

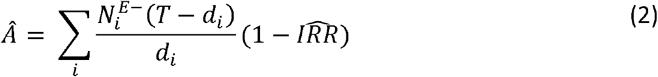

We obtained 95% confidence intervals for *A* using bootstrap (10,000 resampling). For each resampling, a value of *IRR* was randomly sampled based on the parameters estimated in the SCCS analysis.

Finally, based on *Â* and *N*, the total number of outbreaks observed, we obtained the outbreaks prevented fraction, *PF*, with:

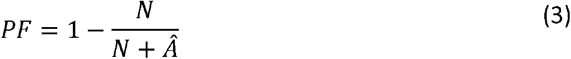

## RESULTS

### Outbreak occurrence and PMVCs

A total of 96 outbreaks out of 97 records (99.0%) were geographically resolved. Among the 479 provinces within the African endemic or at-risk region for yellow fever, 81 (16.9%) from 18 countries (of 34 countries) experienced at least one yellow fever outbreak between 2005 and 2018 (Figure 1A), including 12 provinces experiencing more than one outbreak. The Poisson probability distribution applied satisfactorily to the observed outbreaks distribution (Supplementary Table S1). Overall, 124 (25.9%) provinces were targeted for at least one PMVC (Figure 1B). The SCCS study sample was constituted from 33 (6.9%) provinces having experienced both outbreak and PMVC implementation over the study period. Temporal trends in the estimated population-level vaccination coverage for this sample are displayed in supplementary material (Supplementary Figure S1). The median of the difference between the post- and the pre-PMVC estimate of vaccination coverage was 24.2% (inter-quartile range: 9.4 – 42.7%) (Supplementary Figure S2).

**Figure 1:**
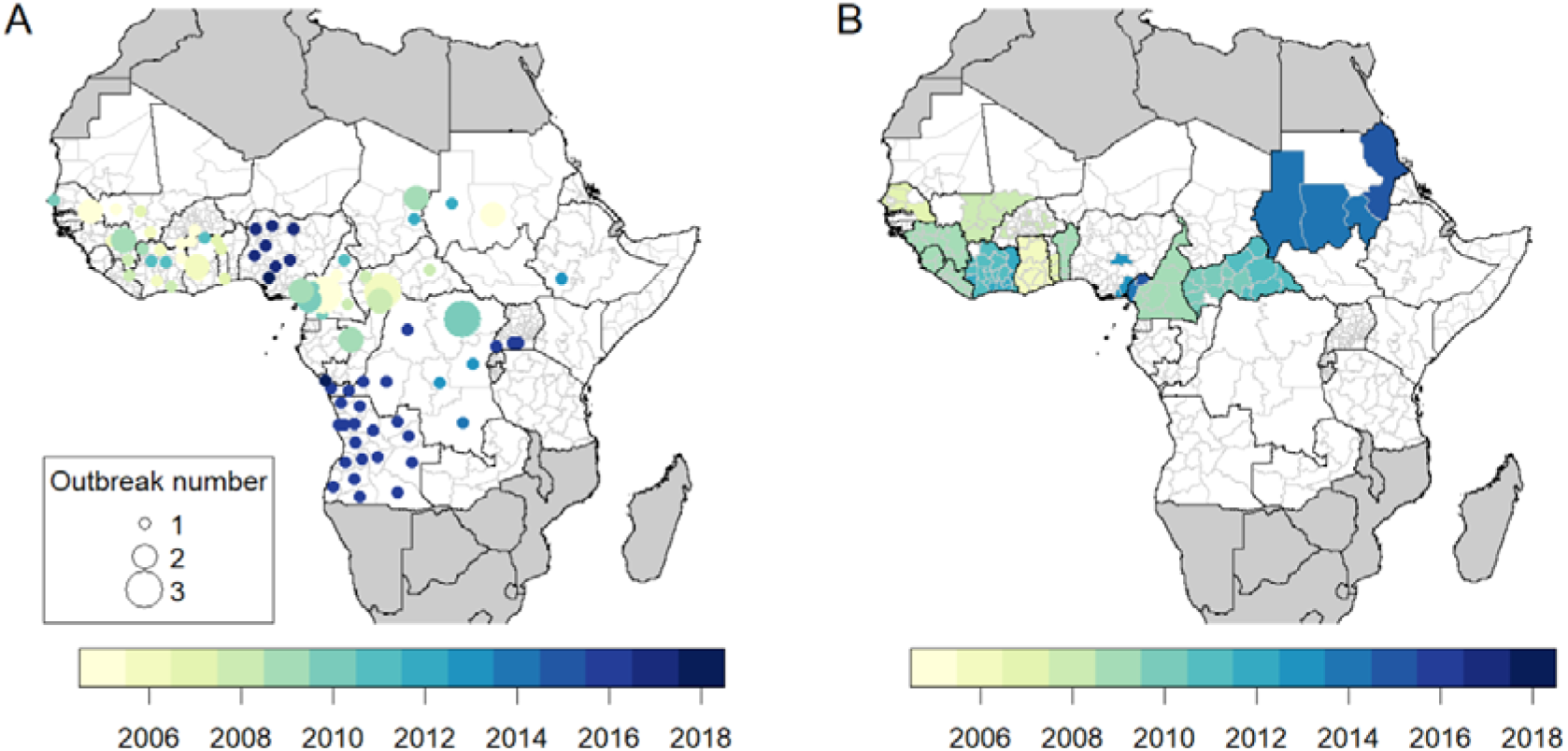
Occurrence of yellow fever outbreaks (A) and preventive mass vaccination campaigns (B) at the province level over the 2005-2018 period. Maps were produced from GADM version 2.0.

### SCCS analysis

Among the SCCS study sample, the first outbreak occurred during the unexposed period in 26 (78.8%) provinces versus 7 (21.2%) occurring in the exposure period (Figure 2). Under baseline assumptions, this corresponded to a significantly reduced incidence rate ratio of 0.14 (95% Confidence interval, CI: 0.06 – 0.34) for the exposed versus unexposed periods. A similar protective association was observed when considering all outbreaks instead of the first one only (IRR = 0.19, 95% CI 0.09 – 0.39) or when including a 3-year pre-exposure period (IRR = 0.14, 95% CI 0.05 – 0.40).

**Figure 2:**
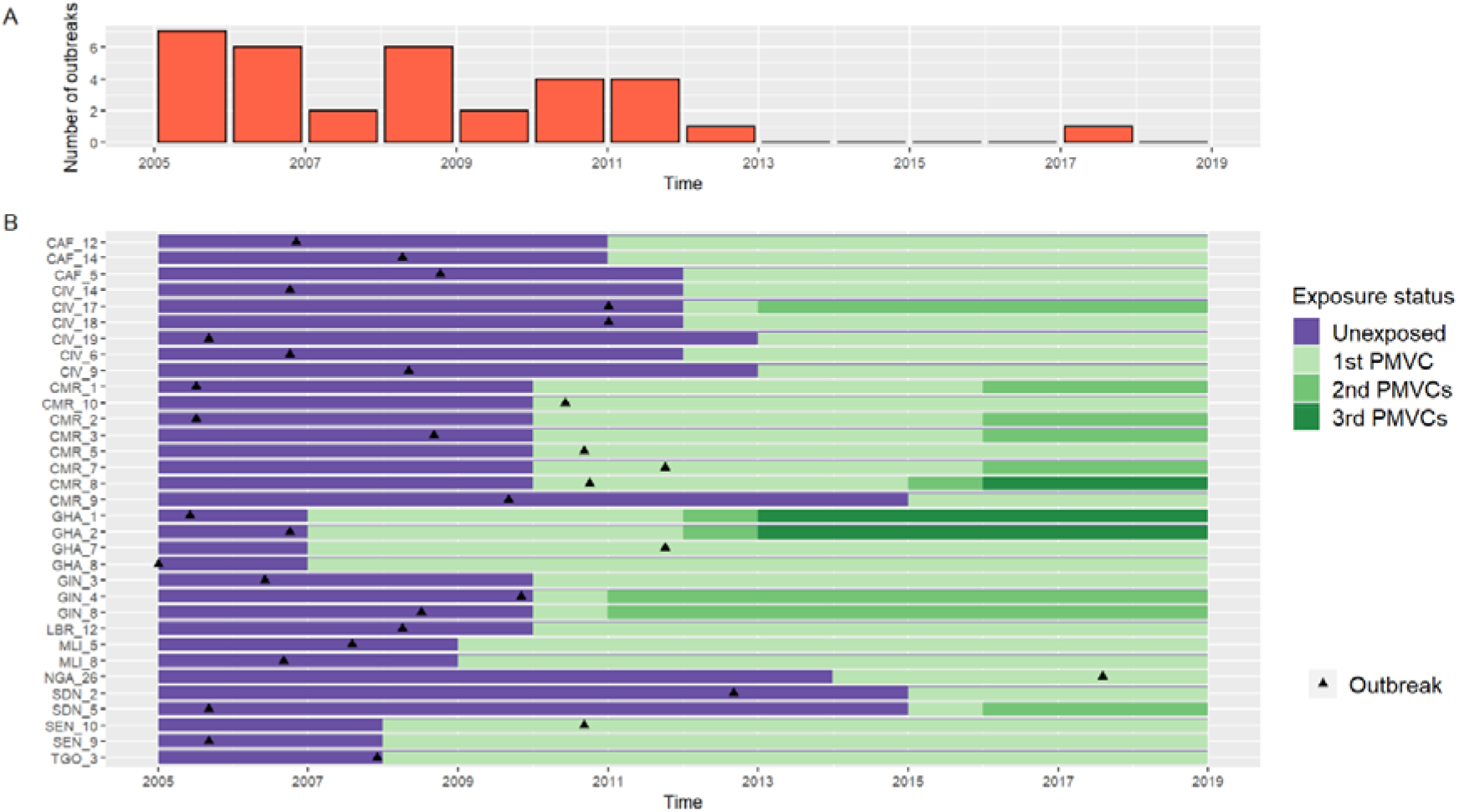
Swimmer plot of the chronology of exposure to preventive mass vaccination campaigns (PMVCs) and yellow fever outbreaks among the 33 exposed cases African provinces (2005-2018). A: Time distribution of yellow fever outbreaks. B : Swimmer plot. The 3-digit code on the y-axis refers to countries ISO codes (see Supplementary Table S2 for complete province and country names).

Considering estimates of population-level vaccine coverage as a categorical variable based on 20% allowed observing a reduced risk of outbreak for higher levels of coverage (Table 2). Considering vaccine coverage as a continuous linear exposure ensured a better fit of the model (likelihood ratio test: p =0.44). Doing so, we estimated that a 10%-increase in vaccine coverage was associated with a risk of outbreak decreased by 41% (IRR 0.59; 95% CI 0.46 – 0.76).

**Table 2:**
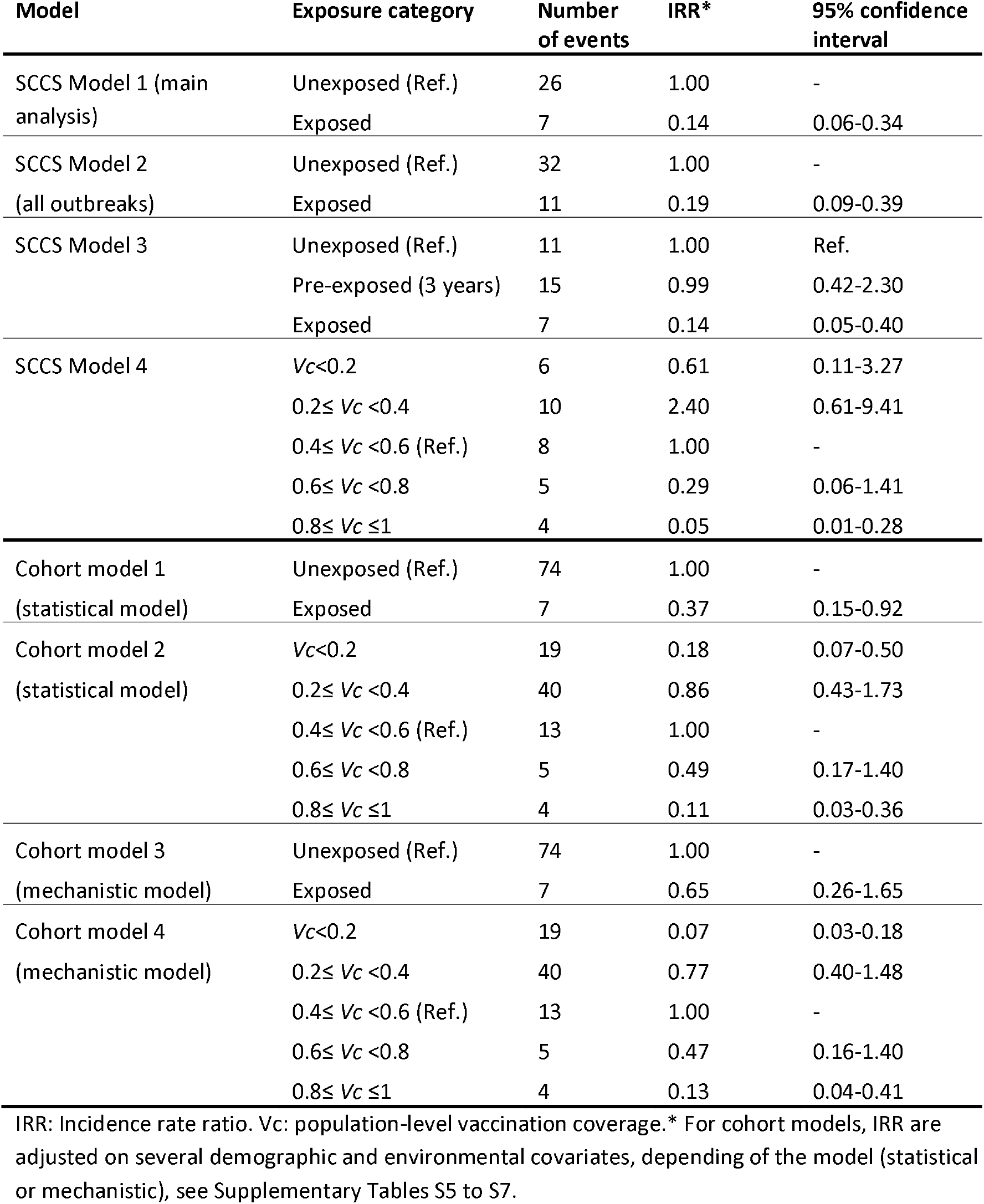
Association between exposure to preventive mass vaccination campaigns (PMVCs) and yellow fever outbreak in African provinces, 2005-2018.

| Model | Exposure category | Number of events | IRR* | 95% confidence interval |
| --- | --- | --- | --- | --- |
| SCCS Model 1 (main analysis) | Unexposed (Ref.) | 26 | 1.00 | - |
|  | Exposed | 7 | 0.14 | 0.06-0.34 |
| SCCS Model 2 (all outbreaks) | Unexposed (Ref.) | 32 | 1.00 | - |
|  | Exposed | 11 | 0.19 | 0.09-0.39 |
| SCCS Model 3 | Unexposed (Ref.) | 11 | 1.00 | Ref. |
|  | Pre-exposed (3 years) | 15 | 0.99 | 0.42-2.30 |
|  | Exposed | 7 | 0.14 | 0.05-0.40 |
| SCCS Model 4 | $V_c < 0.2$ | 6 | 0.61 | 0.11-3.27 |
| | $0.2 \leq V_c < 0.4$ | 10 | 2.40 | 0.61-9.41 |
| | $0.4 \leq V_c < 0.6$ (Ref.) | 8 | 1.00 | - |
| | $0.6 \leq V_c < 0.8$ | 5 | 0.29 | 0.06-1.41 |
| | $0.8 \leq V_c \leq 1$ | 4 | 0.05 | 0.01-0.28 |
| Cohort model 1 (statistical model) | Unexposed (Ref.) | 74 | 1.00 | - |
|  | Exposed | 7 | 0.37 | 0.15-0.92 |
| Cohort model 2 (statistical model) | $V_c < 0.2$ | 19 | 0.18 | 0.07-0.50 |
| | $0.2 \leq V_c < 0.4$ | 40 | 0.86 | 0.43-1.73 |
| | $0.4 \leq V_c < 0.6$ (Ref.) | 13 | 1.00 | - |
| | $0.6 \leq V_c < 0.8$ | 5 | 0.49 | 0.17-1.40 |
| | $0.8 \leq V_c \leq 1$ | 4 | 0.11 | 0.03-0.36 |
| Cohort model 3 (mechanistic model) | Unexposed (Ref.) | 74 | 1.00 | - |
|  | Exposed | 7 | 0.65 | 0.26-1.65 |
| Cohort model 4 (mechanistic model) | $V_c < 0.2$ | 19 | 0.07 | 0.03-0.18 |
| | $0.2 \leq V_c < 0.4$ | 40 | 0.77 | 0.40-1.48 |
| | $0.4 \leq V_c < 0.6$ (Ref.) | 13 | 1.00 | - |
| | $0.6 \leq V_c < 0.8$ | 5 | 0.47 | 0.16-1.40 |
| | $0.8 \leq V_c \leq 1$ | 4 | 0.13 | 0.04-0.41 |

IRR: Incidence rate ratio. $V_c$ : population-level vaccination coverage.\* For cohort models, IRR are adjusted on several demographic and environmental covariates, depending of the model (statistical or mechanistic), see Supplementary Tables S5 to S7.

### Sensitivity analysis

The negative association between exposure to PMVCs and outbreak remained significant across a range of assumptions regarding the imputation of the date (within the same year) of PMVCs implementation and outbreak starting date (when missing), and across various observation periods (Supplementary Table S3 and S4).

When re-sampling 100 times the SCCS study sample while allowing only one sampled province per country, and after excluding re-sampling yielding to random zero in the corresponding contingency table (N=16 with no outbreak occurring during exposed periods, thus leading to infinite confidence interval surrounding the association measure), we obtained an averaged IRR of 0.09 (95% CI 0.01-0.62).

### Cohort-style analysis

In a cohort design, over the 81 outbreaks (first outbreaks only) that occurred over the study period, 74 occurred during unexposed periods versus 7 occurring in exposed periods. Most of the environmental covariates we explored were associated with exposure to PMVCs (Supplementary Table S5). Exposure to PMVCs was associated to a significant reduced risk of outbreak (IRR = 0.37, 95% CI 0.15-0.92) when adjusting on the covariates obtained from a statistical model. When adjusting on covariates obtained from a mechanistic model, exposure to PMVC was not significantly associated with the risk of outbreak (IRR=0.65, 95% CI 0.26-1.65) (detailed results in Supplementary Tables S6 and S7). For both sets of covariates, we observed an inversed U-shaped association between the estimates of vaccination coverage and the risk of outbreak, with the risk decreasing for lowest and highest values of vaccination coverage (Table 2).

### Number of outbreaks averted and prevented fraction

Based on the value of IRR estimated in the main analysis (IRR = 0.14, 95% CI 0.06 – 0.34), we estimated that PMVCs implemented over the study period averted in median 50 (28 to 80) outbreaks. When considering a total of 96 outbreaks occurring over the considered time period, this corresponds to a prevented fraction of 34% (22% to 45%).

## DISCUSSION

In this paper, using the SCCS method, we quantified the preventive effect of PMVCs on the risk of outbreak at the province level, documenting a 86% (95% CI 66 to 94%) reduction of the risk of outbreak occurrence for provinces that were targeted by a PMVC. This result was robust over a range of assumptions. When using an estimate of population-level coverage as exposure, we also observed a dose-response preventive effect on the risk of outbreak. Considering the scale of PMVCs implementation during the study period, this corresponded to an estimated 22% to 45% of outbreaks averted by PMVCs in Africa between 2005 and 2018. Based on a cohort design analysis, the association between PMVC and outbreak was sensitive to the choice of adjustment variables. Moreover, we observed a challenging U-shape association between vaccination coverage and the risk of outbreak in the cohort analysis. Overall, these results suggest a risk of residual confounding that the SCCS method, but not cohort design, could overcome, at least for time-independent confounder. To our knowledge, this is the first time a SCCS analysis was conducted at the population level.

Considering evidence of yellow fever vaccine efficacy at the individual level, a preventive effect of PMVC on outbreak risk was indeed expected. This is why we think that the cohort analysis results may be biased by residual confounding ; whereas we consider the results obtained from the SCCS method to be more trustworthy. Indeed, the indication of provinces for PMVCs partly relies on a risk assessment [3]. For the results of the cohort-design analysis to be valid, one needs to account for all possible confounders in the association between PMVCs and outbreak. This is particularly challenging as the environmental and demographic drivers of yellow fever are not fully understood yet [9].

Another result suggesting residual confounding in the cohort-design analysis is the U-shaped relationship between vaccination coverage and outbreak risk. The yellow fever vaccine has not been introduced in large regions of Eastern Africa yet, as the risk of yellow fever is usually considered as low, though existing (eg. Kenya). This setting can actually yield to a spurious negative association between low level of vaccination coverage and outbreak risk when confounders are not controlled for. In the SCCS analysis, we did observe a linear relationship in the expected association between vaccination coverage and outbreak risk, which stands as another evidence for reduced residual confounding as compared to the cohort analysis. Analysing cases only, instead of the corresponding complete cohort, translates into a loss of efficiency, but previous work showed that this loss is small, especially when the fraction of the sample experiencing the exposure is high [26]. Moreover, this loss of efficiency has to be weighed against a better control of time-invariant confounders. Previous examples illustrated that the SCCS design is likely to produce more trustful results than the corresponding cohort analysis, especially when a strong indication bias is likely [27,28].

Although the SCCS method has been originally developed to be conducted at the individual level, we ensured that our analysis complied with all its requirements [12,15]. Exposure and outcomes were ascertained independently. The list of PMVCs was compiled based on information provided by international funders. Outbreak occurrence were compiled from WHO sources, which themselves compile outbreak notification from countries as per the 2005 International Health Regulation. The observation period was chosen in order to maximize the chance that cases experienced the exposure period. Indeed, our observation period started few times before the launch of the first Yellow Fever initiative, which boosted the use of PMVCs that have been very rare since the 1960s [4,19]. The choice of the long and unlimited exposure period was based on evidence regarding the long-lasting protection conferred by the yellow fever vaccine, and the SCCS method has been previously used successfully while considering long and unlimited risk periods [29].The application of the SCCS design at the population level certainly deserves further methodological assessments to ensure its robustness to specific issues when transposing it at the population level, especially those related to over-dispersed or auto-correlated events. We hope that the present work will stimulate further studies characterizing the advantage and drawbacks of SCCS as compared to more classically used population-level designs, for instance interrupted time-series.

Under the assumption of causality, the incidence risk ratio we estimate represents the average effect for a province of being targeted by a PMVC, which corresponds to the average treatment effect in the counterfactual framework. This average effect is likely to mask large heterogeneity in the local effect of PMVC. Indeed, PMVCs occur in population with various baseline levels of immunity, and they may achieve various levels of post-intervention coverage. The dose-response relationship we observed in the association between vaccination coverage and outbreak risk brought additional evidence for a causal link between PMVC and reduced outbreak risk. When looking at higher values of vaccination coverage, it is notable that several outbreaks (n=4) occurred at estimated levels of vaccination coverage >80%, an empirical threshold that has been often suggested as protecting from outbreaks [30]. While keeping in mind all the limitations such province-based estimates of vaccination coverage may have (outbreaks could occur in small pockets with low vaccination coverage even in provinces with high coverage), this can be viewed an argument to ensure high vaccination coverages homogeneously in at-risk areas, and to sustain them after PMVCs by ensuring routine infant vaccination.

Relying on our estimate of the preventive effect of PMVC, the timing of implementation of these PMVCs and the number outbreaks observed during the study period, we further estimated that PMVCs have averted from 28 to 80 outbreaks in Africa between 2005 and 2018, corresponding to a prevented fraction lying between 22% to 45%. Garske et al. previously estimated that vaccination campaigns conducted up to 2013 averted between 22 to 31% of yellow fever cases and deaths in Africa, while Shearer et al estimated that all vaccination activities (including routine infant vaccination) conducted up to 2016 have averted 33 to 39% of cases [4,5]. Our estimates were in a comparable range, although direct comparison with these model-based estimates is not straightforward. Indeed the latter are expressed as proportions of all yellow fever cases, including sylvatic cases that are not linked to outbreaks. Preventing outbreaks of epidemic prone diseases is critical for ensuring global health security yet there are few empirical studies that quantify the impact of public health interventions like immunization have on the risk of outbreaks.

A main limitation of our study is that it does not account for possible time-varying confounders. Environmental changes affecting vector-borne diseases have been documented across tropical Africa over the study period, probably the main being changing land-use such as deforestation [31,32]. More frequent intrusions of humans into forest and jungles, together with increasing human mobility between endemic and non-endemic areas, have also been suggested to have affected the yellow fever risk in the recent period [33]. Similarly, recent international emphasis about yellow fever may have led to better surveillance of the disease in the recent years. However, these various factors are likely to have increased the risk of outbreaks and the probability of outbreak detection in the recent period, which overlaps with the post-PMVCs period in our study sample. This may have led to an underestimate of the association between PMVC and yellow fever outbreaks. Lastly, historical vaccination activities that occurred up to the 1970s may potentially act as a time-varying confounder. Indeed, the contribution of older people (those potentially exposed to these historical campaigns) to the population-level immunity may decrease over time by population renewal. However, the corresponding bias is likely limited considering the population structure skewed towards younger individuals in the region considered here. Moreover, such a bias is likely to have led to an under-evaluation of the association between PMVC and yellow fever outbreaks. Indeed, decreasing population-level immunity would have increased the risk of outbreak during the more-recent period, which corresponds to the post-PMVCs period. Previous quantification of the outstanding health impact of vaccination activities have mainly focused on cases or deaths prevented while relying on mathematical models, which structures and assumptions may be difficult to understand by a non-expert audience, whether it be decision-makers or targeted populations [34,35]. Here we further document this impact using an empirical, maybe more intuitive approach thus allowing for a triangulation of methods to further document the beneficial impact of yellow fever vaccine campaigns. This method relies on data that are quite easily accessible. Thus, our method could be applied to other diseases for which PMVCs are implemented, such as polio, meningitis or cholera. Due to the COVID-19 pandemic, WHO recommended to temporarily suspend preventive campaigns while assessments of risk, and effective measures for reducing COVID-19 circulation were established. In consequence, regarding yellow fever specifically, four countries postponed vaccination campaigns [36]. Our results provide additional evidence to encourage a rapid rescheduling of these vaccine campaigns in order to prevent further outbreaks of preventable disease.

## Data Availability

All data and codes used for the analysis are publicly available at https://github.com/kjean/YF_outbreak_PMVC

https://github.com/kjean/YF_outbreak_PMVC

## Acknowledgment

We are thankful to Paddy Farrington for helpful discussions.

## REFERENCES

1. World Health Organization. Yellow fever in Africa and the Americas, 2018. Wkly Epidemiol Rec. 2019;94: 365–380.

2. WHO Disease Outbreak News (DONs). [cited 3 Jul 2020]. Available: https://www.who.int/csr/don/archive/disease/yellow_fever/en/

3. World Health Organization. Eliminate Yellow fever Epidemics (EYE): a global strategy, 2017–2026. Wkly Epidemiol Rec. 2017;92: 193–204.

4. Garske T, Van Kerkhove MD, Yactayo S, Ronveaux O, Lewis RF, Staples JE, et al. Yellow Fever in Africa: estimating the burden of disease and impact of mass vaccination from outbreak and serological data. PLoS Med. 2014;11: e1001638. doi:10.1371/journal.pmed.1001638

5. Shearer FM, Longbottom J, Browne AJ, Pigott DM, Brady OJ, Kraemer MUG, et al. Existing and potential infection risk zones of yellow fever worldwide: a modelling analysis. The Lancet Global Health. 2018;0. doi:10.1016/S2214-109X(18)30024-X

6. Jean K, Hamlet A, Benzler J, Cibrelus L, Gaythorpe KAM, Sall A, et al. Eliminating yellow fever epidemics in Africa: Vaccine demand forecast and impact modelling. PLOS Neglected Tropical Diseases. 2020;14: e0008304. doi:10.1371/journal.pntd.0008304

7. WHO Global Malaria Programme. Guidance on temporary malaria control measures in Ebola-affected countrie. 2014. Available: https://apps.who.int/iris/bitstream/handle/10665/141493/WHO_HTM_GMP_2014.10_eng.pdf;jsessionid=8F680505C5FC344148A5579ECB07BC40?sequence=1

8. Walker PGT, White MT, Griffin JT, Reynolds A, Ferguson NM, Ghani AC. Malaria morbidity and mortality in Ebola-affected countries caused by decreased health-care capacity, and the potential effect of mitigation strategies: a modelling analysis. Lancet Infect Dis. 2015;15: 825– 832. doi:10.1016/S1473-3099(15)70124-6

9. Jentes ES, Poumerol G, Gershman MD, Hill DR, Lemarchand J, Lewis RF, et al. The revised global yellow fever risk map and recommendations for vaccination, 2010: consensus of the Informal WHO Working Group on Geographic Risk for Yellow Fever. The Lancet Infectious Diseases. 2011;11: 622–632. doi:10.1016/S1473-3099(11)70147-5

10. Hamlet A, Jean K, Perea W, Yactayo S, Biey J, Kerkhove MV, et al. The seasonal influence of climate and environment on yellow fever transmission across Africa. PLOS Neglected Tropical Diseases. 2018;12: e0006284. doi:10.1371/journal.pntd.0006284

11. Monath TP, Vasconcelos PFC. Yellow fever. Journal of Clinical Virology. 2015;64: 160–173. doi:10.1016/j.jcv.2014.08.030

12. Petersen I, Douglas I, Whitaker H. Self controlled case series methods: an alternative to standard epidemiological study designs. BMJ. 2016;354: i4515. doi:10.1136/bmj.i4515

13. Farrington CP. Relative incidence estimation from case series for vaccine safety evaluation. Biometrics. 1995;51: 228–235.

14. Jean K, Donnelly CA, Ferguson NM, Garske T. A Meta-Analysis of Serological Response Associated with Yellow Fever Vaccination. The American Journal of Tropical Medicine and Hygiene. 2016;95: 1435–1439. doi:10.4269/ajtmh.16-0401

15. Whitaker HJ, Farrington CP, Spiessens B, Musonda P. Tutorial in biostatistics: the self-controlled case series method. Stat Med. 2006;25: 1768–1797. doi:10.1002/sim.2302

16. World Health Organization. Weekly Epidemiological Record (WER). World Health Organization; Available: https://www.who.int/wer/en/

17. World Health Organization. Disease Outbreak News (DON). World Health Organization; Available: https://www.who.int/csr/don/en/

18. World Health Organization. Yellow fever surveillance and outbreak response: revision of case definitions, October 2010. Wkly Epidemiol Rec. 2010;85: 465–472.

19. World Health Organization. The Yellow fever initiative: an introduction. 2016. Available: https://www.who.int/csr/disease/yellowfev/introduction/en/

20. Gotuzzo E, Yactayo S, Córdova E. Efficacy and duration of immunity after yellow fever vaccination: systematic review on the need for a booster every 10 years. Am J Trop Med Hyg. 2013;89: 434–444. doi:10.4269/ajtmh.13-0264

21. Vannice K, Wilder-Smith A, Hombach J. Fractional-Dose Yellow Fever Vaccination — Advancing the Evidence Base. New England Journal of Medicine. 2018;379: 603–605. doi:10.1056/NEJMp1803433

22. Hamlet A, Jean K, Yactayo S, Benzler J, Cibrelus L, Ferguson N, et al. POLICI: A web application for visualising and extracting yellow fever vaccination coverage in Africa. Vaccine. 2019;37: 1384–1388. doi:10.1016/j.vaccine.2019.01.074

23. Simpson SE. A positive event dependence model for self-controlled case series with applications in postmarketing surveillance. Biometrics. 2013;69: 128–136. doi:10.1111/j.1541-0420.2012.01795.x

24. World Health Organization. Yellow fever in Africa and the Americas, 2015. Wkly Epidemiol Rec. 2016;91: 381–388.

25. Dormann CF, Schymanski SJ, Cabral J, Chuine I, Graham C, Hartig F, et al. Correlation and process in species distribution models: bridging a dichotomy. Journal of Biogeography. 2012;39: 2119–2131. doi:10.1111/j.1365-2699.2011.02659.x

26. Whitaker HJ, Hocine MN, Farrington CP. The methodology of self-controlled case series studies. Stat Methods Med Res. 2009;18: 7–26. doi:10.1177/0962280208092342

27. Kramarz P, DeStefano F, Gargiullo PM, Davis RL, Chen RT, Mullooly JP, et al. Does influenza vaccination exacerbate asthma? Analysis of a large cohort of children with asthma. Vaccine Safety Datalink Team. Arch Fam Med. 2000;9: 617–623. doi:10.1001/archfami.9.7.617

28. Douglas IJ, Evans SJW, Hingorani AD, Grosso AM, Timmis A, Hemingway H, et al. Clopidogrel and interaction with proton pump inhibitors: comparison between cohort and within person study designs. BMJ. 2012;345. doi:10.1136/bmj.e4388

29. Taylor B, Miller E, Farrington CP, Petropoulos MC, Favot-Mayaud I, Li J, et al. Autism and measles, mumps, and rubella vaccine: no epidemiological evidence for a causal association. Lancet. 1999;353: 2026–2029. doi:10.1016/s0140-6736(99)01239-8

30. World Health Organization. Yellow fever key facts. 1 May 2018. Available: http://www.who.int/en/news-room/fact-sheets/detail/yellow-fever

31. Faust CL, McCallum HI, Bloomfield LSP, Gottdenker NL, Gillespie TR, Torney CJ, et al. Pathogen spillover during land conversion. Ecology Letters. 2018;21: 471–483. doi:10.1111/ele.12904

32. Esser HJ, Mögling R, Cleton NB, van der Jeugd H, Sprong H, Stroo A, et al. Risk factors associated with sustained circulation of six zoonotic arboviruses: a systematic review for selection of surveillance sites in non-endemic areas. Parasites & Vectors. 2019;12: 265. doi:10.1186/s13071-019-3515-7

33. World Health Organization, UNICEF, GAVI. A global strategy to Eliminate Yellow fever Epidemics (EYE): 2017–2026. 2017. Available: https://apps.who.int/iris/bitstream/handle/10665/272408/9789241513661-eng.pdf

34. Chang AY, Riumallo-Herl C, Perales NA, Clark S, Clark A, Constenla D, et al. The Equity Impact Vaccines May Have On Averting Deaths And Medical Impoverishment In Developing Countries. Health Aff (Millwood). 2018;37: 316–324. doi:10.1377/hlthaff.2017.0861

35. Li X, Mukandavire C, Cucunubá ZM, Abbas K, Clapham HE, Jit M, et al. Estimating the health impact of vaccination against 10 pathogens in 98 low and middle income countries from 2000 to 2030. medRxiv [preprint]. 2019; 19004358. doi:10.1101/19004358

36. World Health Organization. At least 80 million children under one at risk of diseases such as diphtheria, measles and polio as COVID-19 disrupts routine vaccination efforts, warn Gavi, WHO and UNICEF. 22 May 2020 [cited 9 Jun 2020]. Available: https://www.who.int/news-room/detail/22-05-2020-at-least-80-million-children-under-one-at-risk-of-diseases-such-as-diphtheria-measles-and-polio-as-covid-19-disrupts-routine-vaccination-efforts-warn-gavi-who-and-unicef

