## Supplementary material for "Assessing the impact of preventive mass vaccination campaigns on yellow fever outbreaks in Africa : a population-level self-controlled case-series study"

**Supplementary Text S1 : Cohort models and adjustment**

In a cohort design, the choice of covariates to include is critical to prevent bias due to residual confounding. As no clear consensus has emerged on the demographical and environmental drivers of yellow fever, we considered two (partially overlapping) sets of covariates that were previously used to reproduce the occurrence of yellow fever records in Africa at the province level. The first model is a statistical model reproducing the spatial distribution of yellow fever records with no explicit aims at reproducing underlying biological processes. The second model is a mechanistic model that aimed at reproducing the spatial distribution of the disease while including these processes, here the temperature-dependence of the yellow fever virus cycle. Variables included in each model are presented in Table S1.

| Variable | Data source | Statistical model [1] | Mechanistic model [2] |
| --- | --- | --- | --- |
| **Human population size** (log-transformed) | [3,4] | X | X |
| **Proxy for surveillance quality**: country-level per capita rate of reporting suspected cases of fever and jaundice | Yellow Fever Surveillance Database, surveillance database established by the African Regional Office of WHO | X | X |
| **Longitude** | [5] | X |  |
| **Land cover type** | [6] | X |  |
| **Enhanced Vegetation Index:** optimised remote-sensing measure of vegetation | [7] | X | X |
| **Rainfall** | [8] |  | X |
| **Temperature suitability index** | [2] |  | X |

**Table S1:** Covariates entered for the statistical and mechanistic models used in the cohort-style analysis measuring the association between the implementation of preventive mass vaccination campaign and yellow fever outbreak.

**Supplementary Table S1 : Fit of the Poisson probability distribution to outbreak data**

| **Number of outbreaks** | **0** | **1** | **2** | **3** |
| --- | --- | --- | --- | --- |
| **Observed count** | 398 | 69 | 9 | 3 |
| **Simulated counts** : Median and 95% Confidence interval | 392 (375-408) | 78 (63-95) | 8 (3-14) | 0 (0-2) |

**Supplementary Table S1:** Fit of the Poisson probability distribution to outbreak data.

Based on the entire sample of the 479 provinces, we observed a total of 96 outbreaks over the study period. Under the assumption that these outbreaks are distributed according to a Poisson probability distribution, the corresponding Poisson rate is λ = 96/479. The simulated counts were obtained from 10,000 random realizations of a Poisson process of rate λ = 96/479.

**Supplementary Table S2 : Country and province names, Figure 2 (main text)**

| **Province ISO code** | **Country** | **Province** |
| --- | --- | --- |
| CAF_12 | Central African Republic | Ombella-M'Poko |
| CAF_14 | Central African Republic | Ouham-Pendé |
| CAF_5 | Central African Republic | Haute-Kotto |
| CIV_14 | Côte d'Ivoire | Savanes |
| CIV_17 | Côte d'Ivoire | Vallée du Bandama |
| CIV_18 | Côte d'Ivoire | Worodougou |
| CIV_19 | Côte d'Ivoire | Zanzan |
| CIV_6 | Côte d'Ivoire | Fromager |
| CIV_9 | Côte d'Ivoire | Lagunes |
| CMR_1 | Cameroon | Adamaoua |
| CMR_10 | Cameroon | Sud |
| CMR_2 | Cameroon | Centre |
| CMR_3 | Cameroon | Est |
| CMR_5 | Cameroon | Littoral |
| CMR_7 | Cameroon | Nord |
| CMR_8 | Cameroon | Ouest |
| CMR_9 | Cameroon | Sud-Ouest |
| GHA_1 | Ghana | Ashanti |
| GHA_2 | Ghana | Brong Ahafo |
| GHA_7 | Ghana | Upper East |
| GHA_8 | Ghana | Upper West |
| GIN_3 | Guinea | Faranah |
| GIN_4 | Guinea | Kankan |
| GIN_8 | Guinea | Nzérékoré |
| LBR_12 | Liberia | Nimba |
| MLI_5 | Mali | Koulikoro |
| MLI_8 | Mali | Sikasso |
| NGA_26 | Nigeria | Nassarawa |
| SDN_2 | Sudan | Darfur |
| SDN_5 | Sudan | Kordofan |
| SEN_10 | Senegal | Thiès |
| SEN_9 | Senegal | Tambacounda |
| TGO_3 | Togo | Maritime |

**Supplementary Table S2:** Correspondence table of provinces ISO codes from Figure 2 in the main text and complete country and province names.

**Supplementary Figure S1 : Temporal trends in the estimate of population-level vaccination coverage in the study sample provinces.**

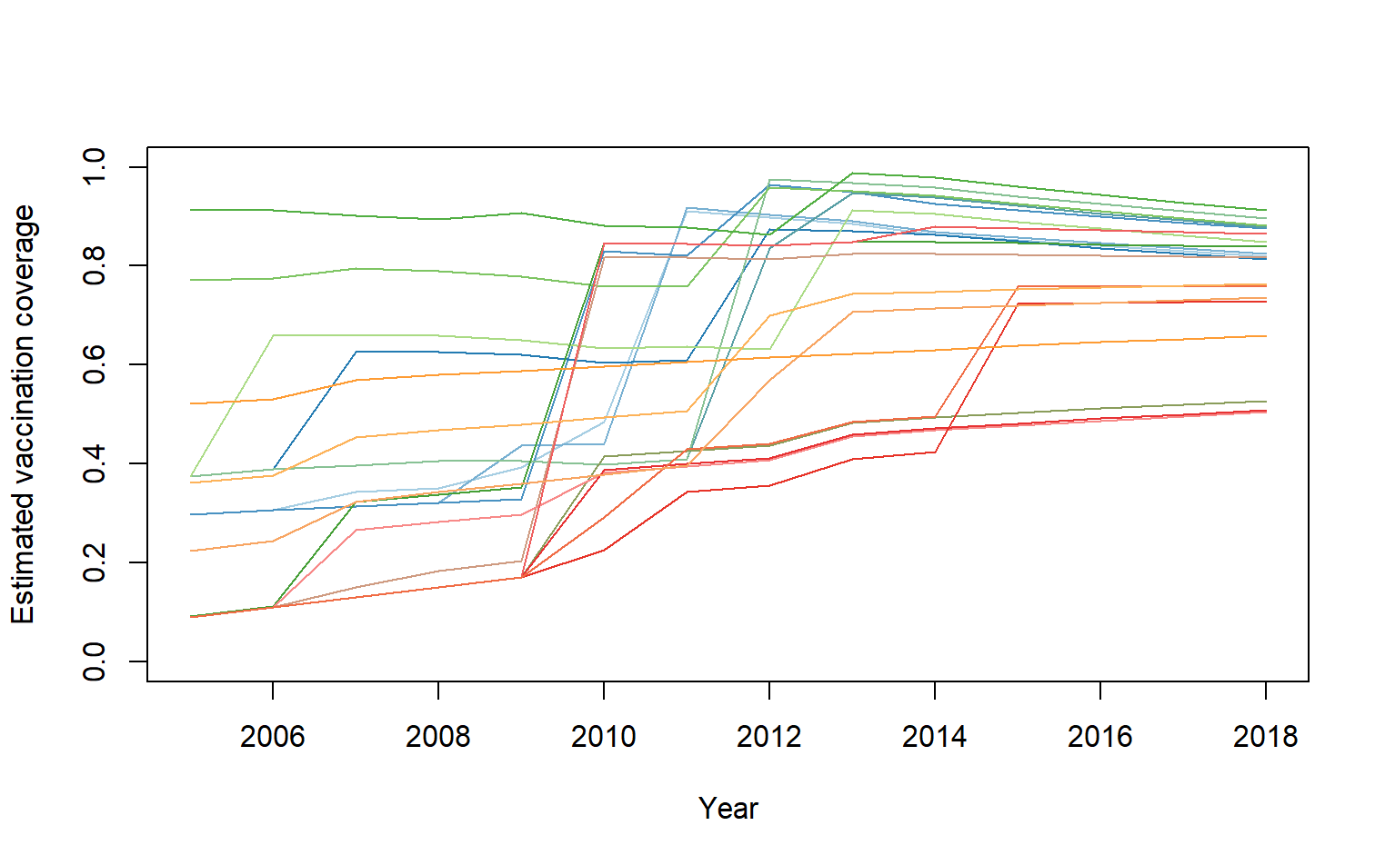

**Figure S1:** Temporal trend in the estimate of population-level vaccination coverage in 33 African provinces having experienced both YF outbreak and the implementation of preventive mass vaccination campaigns over the 2005-2018 study period. Each province is represented by a unique colour.

**Supplementary Figure S2 : Distribution of the difference in the population-level vaccination coverage between the post and the pre-PMVC periods.**

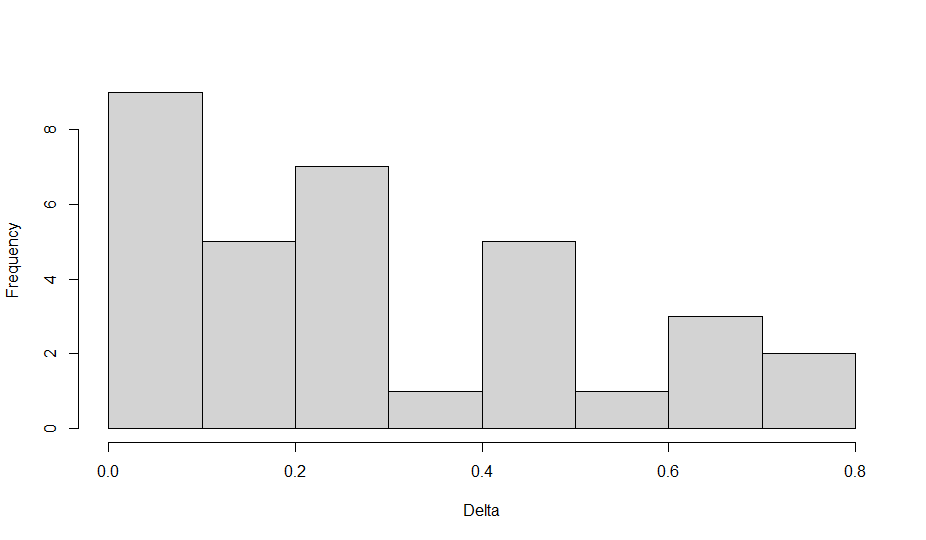
 **Figure S2:** Distribution of the difference in the population-level vaccination coverage between the post and the pre-PMVC periods. PMVC: preventive mass vaccination campaign.

**Supplementary Tables S3 and S4: Sensitivity analysis for the self-controlled case-series method**

| **Model** | **Imputed date of outbreak when missing (within the same year)** | **Imputed date of PMVC when missing (within the same year)** | **Exposure category** | **Number of events** | **IRR*** | **95% confidence interval** |
| --- | --- | --- | --- | --- | --- | --- |
| SCCS Model 1 (main analysis) | July, 1st | Dec, 31st | Unexposed (Ref.)  Exposed | 26  7 | 1.00  0.14 | -  0.06-0.34 |
| Sensitivity analysis #1 | July, 1st | Jan, 1st | Unexposed (Ref.)  Exposed | 23  10 | 1.00  0.16 | -  0.07-0.36 |
| Sensitivity analysis #2 | Dec, 31st | Jan, 1st | Unexposed (Ref.)  Exposed | 21  12 | 1.00  0.22 | -  0.10-0.48 |
| Sensitivity analysis #3 | Jan, 1st | Dec, 31st | Unexposed (Ref.)  Exposed | 26  7 | 1.00  0.14 | -  0.06-0.34 |

**Table S3:** Sensitivity of the self-controlled case-series method results to the imputation of the missing dates of events (outbreak) or exposure (Preventive mass vaccination campaign, PMVC). IRR: incidence rate ratio.

| **Model** | **Beginning of the observation period** | **End of the observation period** | **Exposure category** | **Number of events** | **IRR*** | **95% confidence interval** | **Number of outbreak prevented** | **Total number of outbreak observed on the period** | **Percentage of outbreaks prevented** |
| --- | --- | --- | --- | --- | --- | --- | --- | --- | --- |
| SCCS Model 1 (main analysis) | Jan, 1^st^ 2005 | Dec, 31^st^ 2018 | Unexposed (Ref.)  Exposed | 26  7 | 1.00  0.14 | -  0.06-0.34 | 50 (28 to 80) | 96 | 34% (22% to 45%) |
| Sensitivity analysis #1 | Jan, 1^st^ 2007 | Dec, 31^st^ 2018 | Unexposed (Ref.)  Exposed | 17  7 | 1.00  0.28 | -  0.11-0.70 | 30 (10 to 59) | 79 | 28% (12% to 43%) |
| Sensitivity analysis #2 | Jan, 1^st^ 2005 | Dec, 31^st^ 2014 | Unexposed (Ref.)  Exposed | 26  6 | 1.00  0.12 | -  0.05-0.29 | 24 (10 to 42) | 61 | 29% (14% to 41%) |

**Table S4:** Sensitivity of the self-controlled case-series method results to the choice of start and end dates of the study period. IRR: incidence rate ratio.

**Supplementary Tables S5 to S7 : Univariate and multivariate results of the cohort design analysis**

| **Variable** | **PRR** |
| --- | --- |
| Log population | 0.83 (0.66 - 1.05) |
| Surveillance quality | 1.21 (1.15 - 1.27) |
| Longitude | 0.43 (0.37 - 0.5) |
| Land cover type | 0.60 (0.45 - 0.81) |
| EVI | 1.02 (1.01 - 1.02) |
| Rainfall | 1.23 (1.11 - 1.38) |
| Temperature suitability | 1.48 (1.26 - 1.75) |

**Table S5:** Exposure model: Univariate associations between demographic and environmental variables and implementation of preventive mass vaccination campaigns. PRR: prevalence rate ratio calculated from a modified Poisson regression.

| **Variable** | **uIRR** |
| --- | --- |
| Exposure to PMVC | 0.71 (0.34 – 1.51) |
| Log population | 2.23 (1.54 – 3.22) |
| Surveillance quality | 0.86 (0.69 – 1.07) |
| Longitude | 0.76 (0.65 – 0.90) |
| Land cover type | 0.59 (0.38 – 0.92) |
| EVI | 1.24 (1.03 – 1.50) |
| Rainfall | 1.28 (1.08 – 1.5) |
| Temperature suitability | 0.99 (0.97 – 1.00) |

**Table S6:** Univariate associations between PMVC, demographic and environmental variables and yellow fever outbreak. uIRR: univariate incidence rate ratio.

| **Model** | **Variable** | **aIRR** |
| --- | --- | --- |
| Statistical model (cohort model 1) | Exposure to PMVC | 0.43 (0.18 – 1.02) |
|  | Log population | 3.19 (1.87 – 5.45) |
|  | Surveillance quality | 1.09 (0.88 – 1.34) |
|  | Longitude | 0.63 (0.49 – 0.81) |
|  | Land cover type | 0.44 (0.21 – 0.94) |
|  | EVI | 1.62 (1.28 – 2.03) |
| Mechanistic model (cohort model 3) | Exposure to PMVC | 0.76 (0.32 – 1.81) |
|  | Log population | 2.15 (1.33 – 3.50) |
|  | Surveillance quality | 0.98 (0.77 – 1. 25) |
|  | EVI | 1.25 (0.84 – 1.86) |
|  | Rainfall | 1.17 (0.93 – 1.48) |
|  | Temperature suitability | 1.01 (0.98 – 1.04) |

**Table S7:** Multivariate association between PMVC, demographic and environmental variables and yellow fever outbreak, according to a statistical and a mechanistic model. aIRR: adjusted incidence rate ratio.
